# Preserved Omicron Spike specific antibody binding and Fc-recognition across COVID-19 vaccine platforms

**DOI:** 10.1101/2021.12.24.21268378

**Authors:** Yannic Bartsch, Xin Tong, Jaweon Kang, María José Avendaño, Eileen F. Serrano, Tamara García-Salum, Catalina Pardo-Roa, Arnoldo Riquelme, Rafael A. Medina, Galit Alter

## Abstract

Despite the dramatic spread of Omicron globally, even among highly vaccinated populations, death rates have not increased concomitantly. These data argue that alternative immune mechanisms, beyond neutralization, may continue to confer protection against severe disease. Beyond their ability to bind and block infection, antibodies contribute to control and clearance of multiple infections via their ability to direct antiviral immunity via Fc-effector mechanisms. Thus, here we probed the ability of vaccine induced antibodies, across three COVID-19 vaccines, to drive Fc-effector activity against Omicron. Despite the significant loss of IgM, IgA and IgG binding to the Omicron Receptor Binding Domain (RBD) across BNT162b2, mRNA-1273, and CoronaVac vaccines, stable isotype binding was observed across all of these vaccines to the Omicron Spike. Compromised RBD binding IgG was accompanied by a significant loss of cross RBD-specific antibody Fcγ-receptor binding by the CoronaVac vaccine, but preservation of RBD-specific FcγR2a and Fcγ3a binding across the mRNA vaccines. Conversely, Spike-specific antibodies exhibited persistent binding to Fcγ-receptors, across all three vaccines, albeit higher binding was observed with the mRNA vaccines, marked by a selective preservation of FcγR2a and Fcγ3a binding antibodies. Thus, despite the significant to near complete loss of Omicron neutralization across several vaccine platforms against Omicron, vaccine induced Spike-specific antibodies continue to recognize the virus and recruit Fc-receptors pointing to a persistent capacity for extra-neutralizing antibodies to contribute Omicron disease attenuation.

## Introduction

Antibodies represent the primary correlate of immunity following immunization with nearly all licensed vaccines (*1*), providing protection either via direct blockade of infection or via their ability to leverage the immune system to eliminate pathogens, should the pathogens breach the portal of entry (*2*). Emerging data from SARS-CoV-2 Phase3 vaccine studies clearly demonstrate a critical association between neutralizing and binding antibodies and protection against severe COVID-19 infection(*3*). Yet, the emergence of SARS-CoV-2 variants of concern (VOC), including the Omicron variants, which evade neutralizing antibodies, has led to increased breakthrough infections globally among vaccinated individuals. Thus far, despite this striking rise in breakthrough infections, a concomitant rise in severe disease and death has not been observed, suggesting that vaccine mediated protection may still persist in the setting of a loss of neutralizing antibody activity, pointing to a potential critical role for alternative vaccine induced immune responses as critical modulators of disease severity, the ultimate goal of protection.

Beyond blockade of infection, cellular immune responses can directly or indirectly contribute to protection against severe disease. T cells may directly recognize and eliminate infected cells (*4*). In addition, binding antibodies with the capability of interacting with Fc-receptors (FcRs), found on immune cells, can leverage the antiviral activity of the innate immune system (*5-9*). This drives rapid opsonophagocytic clearance, infected cell cytotoxicity, or pro/anti-inflammatory mediators, etc. each of which have been linked to protection against several viruses including Influenza(*10, 11*), Ebola virus (*12, 13*), HIV (*14*), and most recently against SARS-CoV2 (*6-8*). However, whether Fc-activity persists to provide protection against Omicron, remains unclear. Thus, here we examined whether persistent Fc-activity could partially explain persistent protection against death following Omicron infection. Here we show diminished antibody isotype binding to the Omicron RBD across vaccine platforms, but persistence of robust Fc-activity to the Omicron Spike, which likely contributes to rapidly control and clear viral infection, thereby continuing to attenuate disease severity.

## Results

### Loss of Omicron RBD recognition across vaccine induced immunity

Despite the significant loss of vaccine induced neutralization against the novel Omicron VOC, persistence of vaccine induced antibody binding may continue to confer protection against disease via additional extra-neutralizing antibody functions that have been linked to natural resolution of infection and vaccination (*6-8*). Thus, we probed the persistence of vaccine induced antibody isotype binding to the recombinant receptor binding domain (RBD) across VOCs of the SARS-CoV-2 Spike antigen (Figure 1A). Persistence of RBD recognition was compared using plasma samples from 3 vaccine platforms, including the Moderna mRNA-1273(*15*), Pfizer/BioNtech BNT162b2(*16*), and Sinovac CoronaVac (*17*), all profiled at peak immunogenicity (see methods).

**Figure 1:**
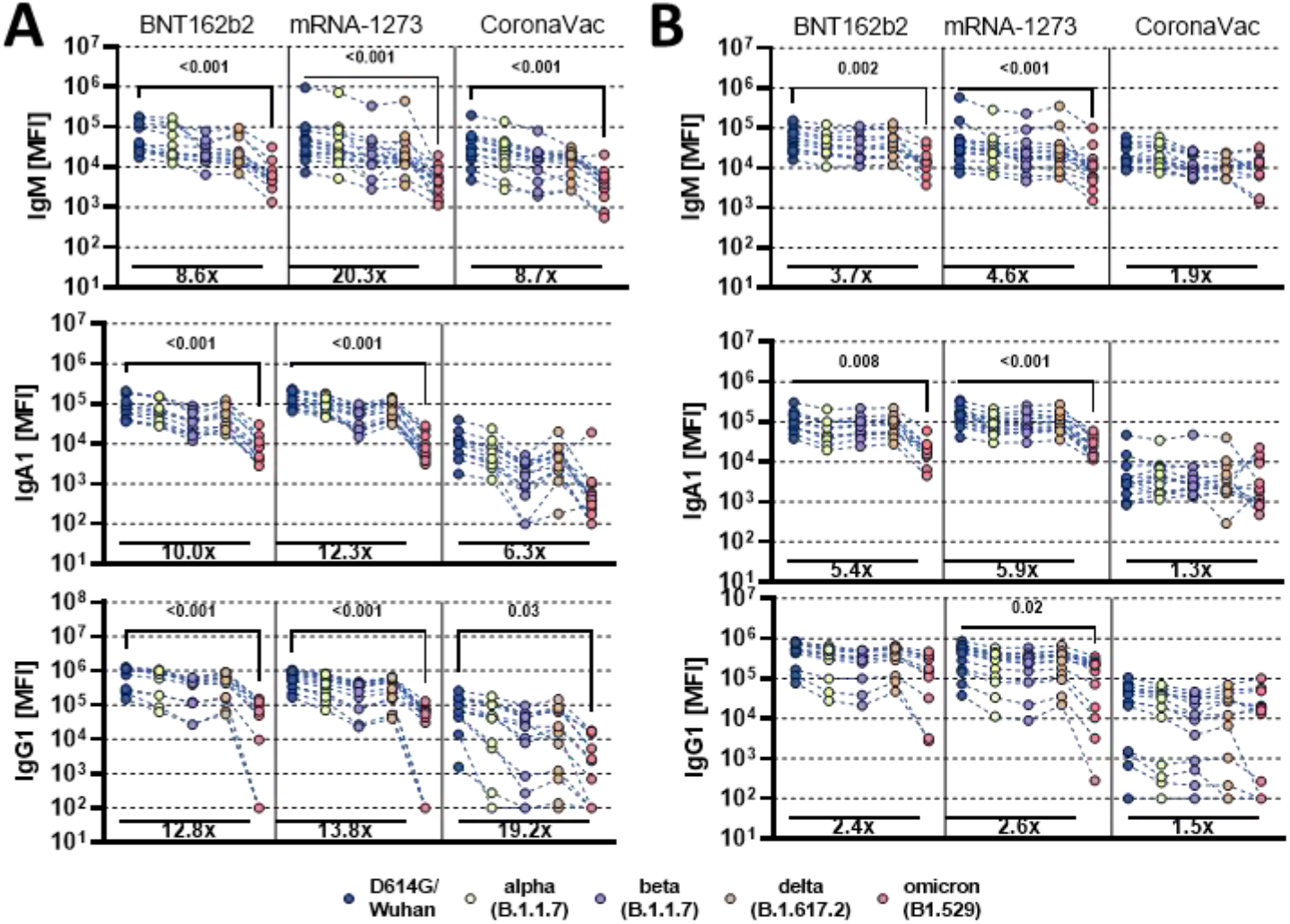
Vaccine induced antibody binding to different SARS-CoV-2 variants of concern. Individuals either received the full dose regimen of the BNT162b2(n = 11), mRNA-1273(n=14), or the aluminum adjuvanted inactivated particle vaccine CoronaVac (n=13). Samples were taken at peak immunogenicity 2 weeks after the last dose. IgM, IgA1 and IgG1 binding titers to D614G (WT), Alpha (B1.117), Beta (B1.351), Delta (B.1.617.2), and Omicron (B1.529) variants of concern receptor binding domain (A) or full Spike (B) were measured by Luminex. Background corrected data is shown and negative values were set to 100 for graphing purposes. A Kruskal-Wallis test with a Benjamini-Hochberg post-test correcting for multiple comparisons was used to test for statistical differences between wildtype variant and omicron titer. P-values for significant different features are shown above and fold change reduction of omicron titer compared to wildtype below each dataset.

Comparable IgM responses were observed across all vaccine platforms to the RBD from D614G (WT), Alpha (B1.117), Beta (B1.351), and Delta (B.1.617.2), with a significant loss of Omicron (B1.529) recognition by all three platforms (**Figure 1A**). mRNA vaccines induced stronger IgA RBD-specific cross-VOC responses, that were compromised, but not completely lost for Omicron. Moderate IgA responses were observed across RBD VOCs following CoronaVac vaccination, that recognized the Beta and Omicron variants to a lesser degree. Robust IgG responses were observed for mRNA vaccines, that were relatively stable across VOC RBDs, including a decrease, but not complete loss of recognition of the Omicron RBD. As expected, CoronaVac induced lower IgG responses across VOC RBDs, that also exhibited diminished binding to the Omicron RBD. These data point to the persistent, albeit lower, recognition of the Omicron RBD across isotypes by specific vaccine platforms.

### Persistent recognition of Omicron Spike across vaccine platforms

While most neutralizing antibodies, that block viral infection, target the Spike antigen on or proximal to the RBD (*18*), Fc-functional antibodies that drive clearance or killing of virus or infected cells can target the whole surface of the Spike antigen. Thus, we next profiled isotype recognition across Spike VOCs (**Figure 1B**). All vaccines induced Spike-specific IgM responses across most VOCs and exhibited attenuated but not significantly reduced binding to the Omicron Spike. Cross-Spike VOC IgA responses were most robustly induced by the BNT162b2 and mRNA-1273 vaccines, but exhibited a partial decline in recognition of the Omicron Spike. Conversely, the CoronaVac vaccine elicited lower IgA responses across Spike VOCs that were completely preserved to Omicron. Moreover, BNT162b2 and mRNA-1273 mRNA vaccines induced the highest levels of cross-Spike VOC IgG binding, which exhibited only a moderate loss of recognition of the Omicron Spike. Interestingly, despite the lower overall IgG titers induced by the CoronaVac vaccine, IgG responses recognized the Omicron spike identically to the wildtype spike, pointing to robust preservation of Spike IgG immunity across the 3 vaccine platforms.

### Variable Omicron-specific Fc-receptor binding activity across vaccine platforms

The ability of antibodies to leverage Fc-effector functions depends on their ability to interact with Fc-receptors (FcR) found on all immune cells (*19*). Thus, we profiled the ability of vaccine induced RBD and Spike-specific antibodies to interact with the four low affinity FcγRs found in humans, known to regulate and drive antibody effector functions (*20*). mRNA vaccines induced robust cross-VOC RBD-specific FcγR binding antibodies but exhibited a near complete loss of inhibitory FcγR2b and neutrophil specific FcγR3b binding, while preserving detectable opsonophagocytic FcγR2a and cytotoxic FcγR3a binding to the Omicron RBD (**Figure 2A**). Conversely, Spike-specific FcR binding persisted more robustly to Omicron (**Figure 2B**). CoronaVac induced intermediate levels of RBD-specific FcγR binding antibodies across VOCs, but exhibited a near complete loss of Omicron RBD-specific FcγR binding, despite the ability to bind to RBD (**Figure 1A**). These data point to qualitative differences in antibody Fc-binding capabilities that are not always linked to antibody titers. Conversely, although CoronaVac induced lower overall IgG levels of Spike-specific antibodies, which recognized the Omicron Spike comparably to the wildtype antigen (**Figure 1B**), CoronaVac Spike antibodies exhibited a more profound decline in Omicron-specific FcγR-binding (**Figure 2A**). However, a common pattern of Omicron Spike-specific FcγR-binding loss was observed across the three platforms, marked by a selective persistence of higher opsonophagocytic FcγR2a and cytotoxic FcγR3a receptor binding, and a sharper decline of inhibitory FcγR2b and neutrophil-activating FcγR3b binding. Thus, despite the significant complete loss of Omicron RBD-specific FcγR binding antibodies, the persistence of robust levels of Omicron Spike-specific FcγR2a and cytotoxic FcγR3a binding antibodies likely may continue to recognize, control, and clear the virus following transmission thereby attenuating disease despite increases in transmission.

**Figure 2:**
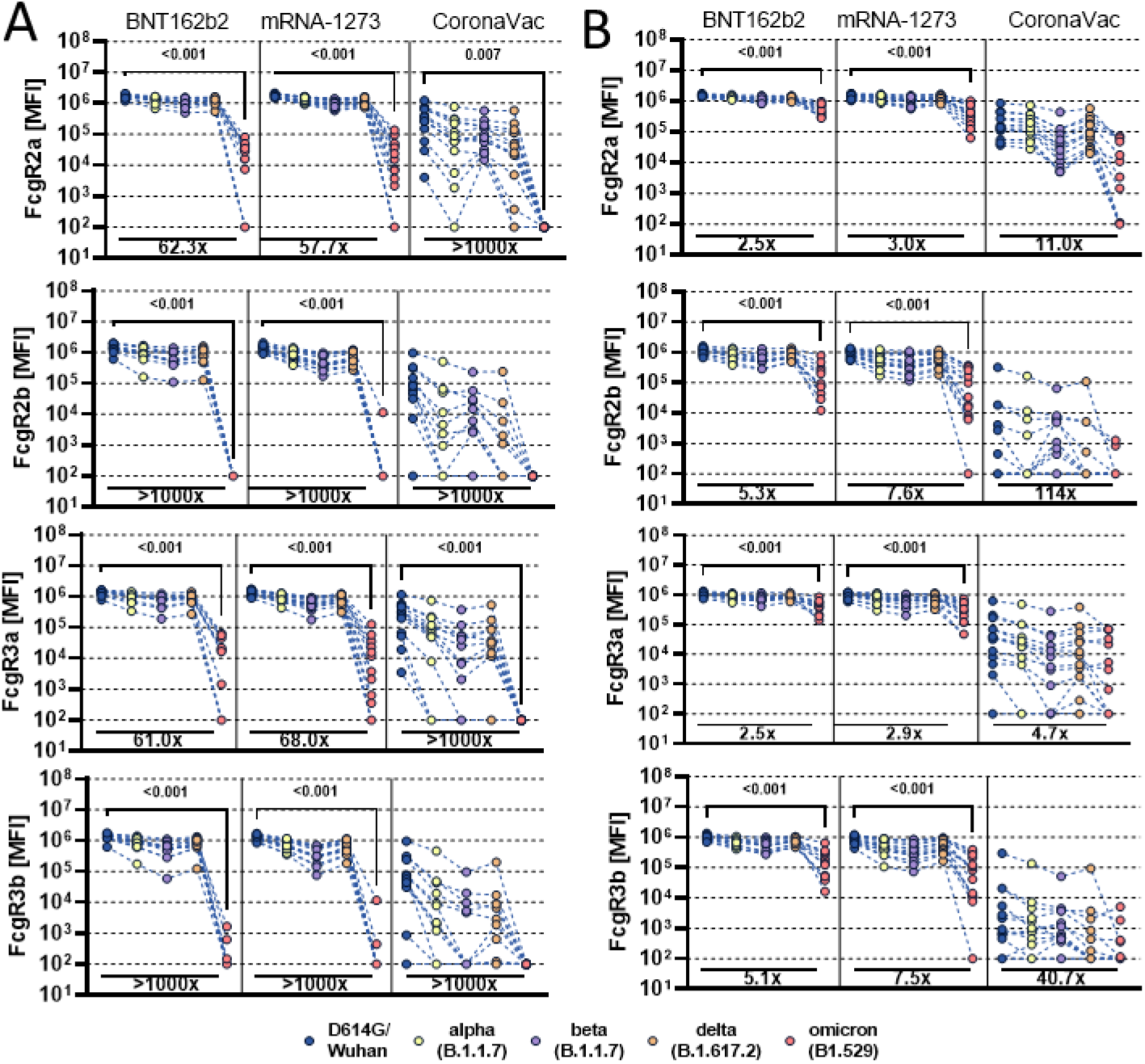
Vaccine induced Fcγ-receptor binding antibody profiles across SARS-CoV-2 variants of concern. Individuals either received the full dose regimen of the BNT162b2(n = 11), mRNA-1273(n=14), or the aluminum adjuvanted inactivated particle vaccine CoronaVac (n=13). Samples were profiled at peak immunogenicity 2 weeks after the last dose. Binding to FcγR2a, FcγR2b, FcγR3a and FcγR3b of D614G (WT), Alpha (B1.117), Beta (B1.351), Delta (B.1.617.2), and Omicron (B1.529) variant of concern receptor binding domain (A) or full Spike (B) specific antibodies were determined by Luminex. Background corrected data is shown and negative values were set to 100 for graphing purposes. A Kruskal-Wallis test with a Benjamini-Hochberg post-test correcting for multiple comparisons was used to test for statistical differences between wildtype variant and omicron titer. P-values for significant different features are shown above and fold change reduction of omicron titer compared to wildtype below each dataset.

## Discussion

As SARS-CoV-2 continues to evolve as it adapts to its new host, the virus has acquired a progressive collection mutations preferentially within the S1 domain of the Spike antigen, within or proximal to the receptor binding domain (RBD), aimed at enhancing Spike binding to the angiotensin-converting enzyme 2 (ACE2) receptor (*21*). Because many of the most potent neutralizing antibodies bind to the RBD, aimed at interfering or blocking interactions with ACE2, both vaccine induced neutralizing antibodies and monoclonal therapeutics have progressively lost neutralization potency against emerging variants of concern (VOC)(*5, 22*). Yet unlike previous VOCs, Omicron possesses more than 40 mutations, including 29 in the Spike protein, that, to date, represents the most profound escape from natural and vaccine induced neutralizing antibody activity. This loss of neutralization, coupled to enhanced ACE2-binding, accounts for the remarkable rise in transmission events globally. However, as a second line defense, following infection, both direct and indirect cellular mechanisms contribute to pathogen control and clearance. Specifically, T cells may directly recognize and kill infected cells (*9*). Additionally, antibodies able to leverage innate immune activity can both drive the rapid elimination of viral particles as well as deploy the cytotoxic power of the immune system to kill infected cells(*19*). While emerging data point to persistent COVID-19 vaccine induced T cell recognition of Omicron (*4*), it was unclear whether vaccine induced antibodies continue to leverage Fc-activity against this novel VOC.

Here we observed a more pronounced loss of Omicron-RBD compared to Omicron-Spike isotype/subclass and FcR binding was observed across vaccine platforms, likely linked to the preferential incorporation of mutations in the S1 domain of the SARS-CoV-2 spike. However, unlike neutralizing antibodies, that must target a limited number of regions on the Spike, involved in attachment, positioning of the RBD, or fusion, antibodies that mediate Fc-activity can likely bind across the entire antigenic surface. Fc-activity solely requires formation of immune complexes and arrangements of antibodies with Fc-domains that are accessible to local immune cells. The persistence of Omicron-IgG binding to the Spike antigen across the mRNA and inactivated vaccine platforms suggests that vaccine induced antibodies may continue to opsonize the virus and virally infected cells even after infection with of Omicron. Thus, while neutralizing antibodies are likely to be key to blocking transmission, non-neutralizing antibodies able to leverage Fc-biology may contribute to persistent disease attenuation.

While the three vaccines maintained more robust binding to Omicron Spike specific FcγR2a, a selective loss of FcγR2b and FcγR3b was observed across the platforms. FcγR2b is the sole low-affinity inhibitory receptor in humans, likely involved in attenuated inflammatory activity (*20*). Likewise, FcγR3b is solely expressed on neutrophils, likely critical for rapid opsonophagocytic clearance of opsonized viral particles (*20*). While continued binding to FcγR2a and FcγR3a may lead to continued clearance of particles and killing of infected cells, the loss of FcγR2b and FcγR3b may result in a more inflammatory response, that may translate to symptoms, but may still attenuate severity and death. Likewise, emerging epidemiological reports suggest that Omicron infection, while less severe, causes mild to moderate symptomatic infection(*23, 24*). However, real-world comparisons of symptom severity across vaccine platforms are needed to provide enhanced resolution of the roles of individuals FcγRs in attenuating disease.

While many developed countries have begun aggressive boosting campaigns, the majority of the world, where many variants may evolve, remains incompletely vaccinated. Thus, understanding the ability of distinct vaccines to drive immunity to Omicron is urgently needed. Moreover, defining the immunological mechanisms that contribute to disease attenuation, in the absence of neutralization, may provide key insights to guide effective pan VOC-vaccine design and boosting campaigns to help control the global COVID-19 pandemic. Here we demonstrate the persistence of Omicron Spike-, but reduced RBD-, specific binding and Fc-activating potential across vaccine platforms, providing some initial insights on persisting mechanisms that may contribute to disease attenuation despite the significant loss of neutralization to this novel SARS-CoV2 variant of concern.

## Data Availability

This study was overseen and approved by the MassGeneral Institutional Review Board (IRB #2020P00955 and #2021P002628).

## Data Availability Statement

All data produced in the present work are contained in the manuscript

## Acknowledgments

We thank Nancy Zimmerman, Mark and Lisa Schwartz, an anonymous donor (financial support), Terry and Susan Ragon, and the SAMANA Kay MGH Research Scholars award for their support. We acknowledge support from the Ragon Institute of MGH, MIT, and Harvard, the Massachusetts Consortium on Pathogen Readiness (MassCPR) and the Musk foundation. This study was funded by the NIH (3R37AI080289-11S1, R01AI146785, U19AI42790-01, U19AI135995-02, U19AI42790-01, 1U01CA260476-01, CIVIC75N93019C00052 and The Gates Foundation Global Health Vaccine Accelerator Platform (OPP1146996 and INV-001650). Work in the Medina Laboratory was based on protocols and the study set-up established in part with the support from FONDECYT 1212023 grant from the Agencia Nacional de Investigación y Desarrollo (ANID) of Chile, the FLUOMICS Consortium (NIH-NIAD grant U19AI135972) and the Center for Research on Influenza Pathogenesis (CRIP), an NIAID Center of Excellence for Influenza Research and Surveillance (CEIRS, contract # HHSN272201400008C). We greatly appreciate the outstanding technical work of Erick Salinas, Estefany Poblete and Andres Muñoz. MJA and ES conducted this work as part of their Ph.D. Thesis, Programa de Doctorado en Ciencias Biológicas mención Genética Molecular y Microbiología, Facultad de Ciencias Biológicas, Pontificia Universidad Cátolica de Chile. This work used samples from the phase 1 mRNA-1273 study (NCT04283461; DOI: 10.1056/NEJMoa2022483). The mRNA-1273 phase 1 study was sponsored and primarily funded by the National Institute of Allergy and Infectious Diseases (NIAID), National Institutes of Health (NIH), Bethesda, MD. This trial has been funded in part with federal funds from the NIAID under grant awards UM1AI148373, to Kaiser Washington; UM1AI148576, UM1AI148684, and NIH P51 OD011132, to Emory University; NIH AID AI149644, and contract award HHSN272201500002C, to Emmes. Funding for the manufacture of mRNA-1273 phase 1 material was provided by the Coalition for Epidemic Preparedness Innovation.

## Methods

### Study population

To compare antibody responses elicited by the different vaccines, samples were obtained at peak immunogenicity timepoints from individuals who were vaccinated with the full dose regimen recommended by the respective manufacturer. As part of a phase 1 clinical trial in the US (NCT04283461) individuals received two doses 100 μg mRNA-1273 at days 0 and 28 and samples taken two weeks after the second dose. BNT162b2 vaccinated individuals received 30 μg BNT162b2 at days 0 and 21 and samples were taken two weeks after the second dose. Individuals from Chile received two doses 600U CoronaVac four weeks apart and samples were taken two weeks after the second dose. For the CoronaVac study informed written consent was obtained under protocol 200829003 which was reviewed and approved by the Scientific Ethics Committee at Pontificia Universidad Católica de Chile (PUC). This study was overseen and approved by the MassGeneral Institutional Review Board (IRB #2020P00955 and #2021P002628).

### Antigens

Receptor-binding domain antigens for the wildtype (Wuhan), alpha (B.1.1.7), beta (B.1.351), and delta (B.1.617.2) VOCs were obtained from Sino-Biologicals. Omicron RBD was generously provided by Moderna Inc. Stabilized (hexa-pro) spike of D614G or respective variants was produced in HEK293 cells.

### IgG subclass, isotype and FcγR binding

Antigen specific antibody subclass and isotypes, and FcγR binding was analyzed by Luminex multiplexing. The antigens were coupled to magnetic Luminex beads (Luminex Corp, TX, USA) by carbodiimide-NHS ester-coupling with an individual region per antigen. Coupled beads were incubated with different plasma dilutions (1:100 for IgG2, IgG3, IgG4, IgM and IgA1, 1:500 for IgG1 and 1:1,000 for FcγR probing) for 2 hours at room temperature in 384 well plates (Greiner Bio-One, Germany). Unbound antibodies were washed away and subclasses, isotypes were detected with a respective PE-conjugated antibody (anti-human IgG1, IgG2, IgG3, IgG4, IgM or IgA1 all SouthernBiotech, AL, USA) at a 1:100 dilution. For the analysis of FcγR binding PE-Streptavidin (Agilent Technologies, CA, USA) was coupled to recombinant and biotinylated human FcγR2a, FcγR2b, FcγR3a, or FcγR3b protein. Coupled FcγR were used as a secondary probe at a 1:1000 dilution. After 1 h incubation, excessive secondary reagent was washed away and the relative antibody concentration per antigen determined on an IQue analyzer (IntelliCyt).

### Statistical analysis

If not stated otherwise, we assumed non-normal distributions and plots were generated and statistical differences between two groups were calculated in Graph Pad Prism V.8. A Kruskal-Wallis test with a Benjamini-Hochberg post-test correcting for multiple comparisons was used to test for statistical differences between wildtype variant and omicron titer.

